# A Proteomic Profile of the Ocular Phenome: Systemic Signatures, Predictive Value, and Causal Insights Across 131 Ocular Diseases and Traits

**DOI:** 10.1101/2025.07.31.25332495

**Authors:** Duanke Liu, Hanruo Liu, Xiaoniao Chen, Xinyuan Zhang, Jingxue Zhang, Jian Wu, Li Chen, Ningli Wang

## Abstract

Vision loss remains one of the most pervasive and preventable global health burdens, yet ophthalmology has yet to fully benefit from molecular precision medicine. Unlike oncology and cardiometabolic diseases, the early detection and subtyping of eye disorders are hindered by limited access to intraocular tissues and a prevailing belief that the blood-ocular barrier precludes systemic biomarker utility. To systematically evaluate the relevance of the plasma proteome to ocular phenotypes, we profiled 2,920 circulating proteins in 53,016 UK Biobank participants across 80 clinically defined eye diseases and 51 quantitative ocular traits. We integrated association analyses, protein-based prediction models, Mendelian randomization, and unsupervised clustering to uncover predictive, causal, and mechanistic insights. We identified >2,700 significant protein-disease and >3,100 protein-trait associations, revealing widespread links between systemic proteins and intraocular features—particularly in diabetic retinopathy, intraocular pressure, and ISOS-RPE thickness. Plasma proteins such as GDF15, VSIG4, and PLAUR were consistently associated across phenotypes, implicating inflammation, vascular leakage, and complement signaling as convergent mechanisms. Proteome-based models outperformed clinical predictors in multiple conditions (e.g., AUC = 0.913 for diabetic retinopathy) and identified distinct risk gradients. Mendelian randomization supported causal roles for 144 proteins, including therapeutically actionable targets. Hierarchical clustering of 80 ocular phenotypes revealed six proteome-defined ocular modules, linking anatomically diverse traits through shared systemic biology and suggesting a new molecular taxonomy of eye health. This study provides the first comprehensive map of systemic protein signatures across the ocular phenome. By revealing biologically coherent associations across anatomically diverse ocular traits, our findings underscore the relevance of circulating proteins in reflecting both local ocular pathology and broader systemic physiology. These insights support the use of plasma proteomics for early detection, disease subtyping, and therapeutic exploration in ophthalmology, and position the eye as a clinically informative site of systemic biological signaling.

**Graphical abstract:** 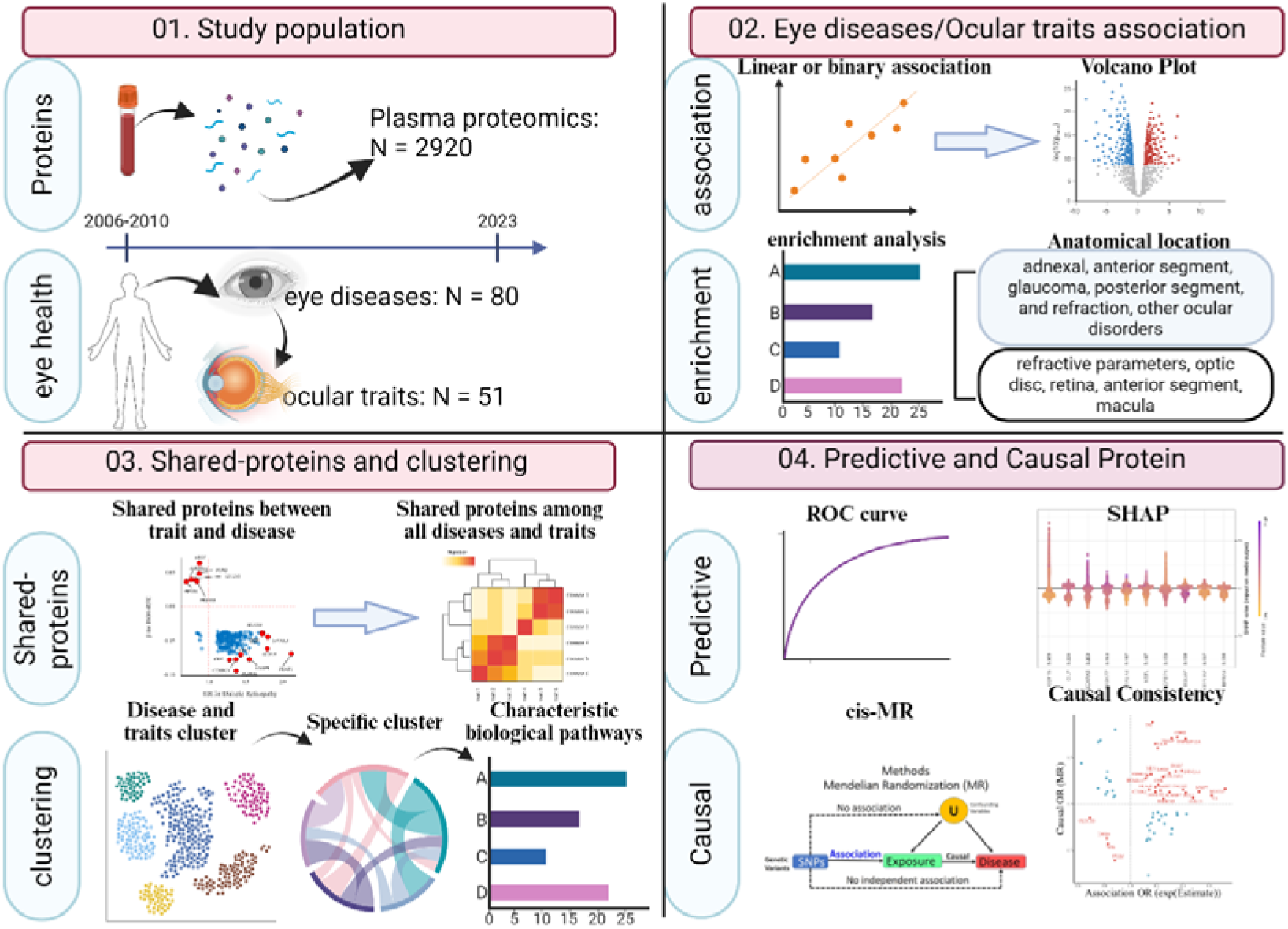

## Introduction

Vision loss affects more than 2.2 billion people globally, representing one of the most pervasive and preventable threats to human health and independence^1^. While up to half of these cases are avoidable, the current landscape of ocular diagnostics remains largely reactive, relying on late-stage imaging and clinical symptoms. A major barrier to earlier intervention lies in the inaccessibility of ocular tissues and fluids, which limits the implementation of molecular profiling strategies that have transformed other areas of medicine^2^ ^3^. Unlike oncology or cardiometabolic disease, ophthalmology has yet to fully enter the era of molecular precision.

Circulating plasma proteins offer an underutilized yet promising window into systemic and organ-specific pathophysiology. Shaped by genetic, environmental, and lifestyle factors, the plasma proteome has been leveraged to uncover biomarkers, enhance risk prediction, and identify therapeutic targets across diverse disease areas^4–9^. However, its application to ocular diseases has remained limited in scope and inconsistent in results. Early studies, often constrained by small sample sizes and narrow phenotypic focus, have reported modest associations between plasma proteins and selected eye conditions, raising concerns about clinical utility. The unique immunological and anatomical features of the eye—particularly the presence of the blood-ocular barrier—have been proposed as biological explanations for this underperformance^6^ ^10–12^.

Yet this perspective may overlook the fundamental nature of many ocular diseases. Conditions such as diabetic retinopathy, age-related macular degeneration, and hypertensive retinopathy are not isolated ocular disorders; rather, they represent local manifestations of systemic metabolic, vascular, and inflammatory disturbances^13–15^. Under pathological conditions, the integrity of the blood-retinal barrier may be disrupted, allowing systemic factors to influence intraocular environments. Such molecular changes may precede clinical diagnosis, offering a powerful yet underexplored opportunity for early prediction and intervention.

Despite this biological rationale, three critical questions remain unanswered. First, do plasma proteins encode shared molecular signatures across the ocular phenome, or are their associations disease- and trait-specific? Second, can circulating proteins offer predictive value for ocular outcomes beyond demographic and clinical variables? Third, are any of these associations driven by causal biology, pointing to modifiable targets for intervention? Addressing these questions requires high-dimensional proteomic data integrated with granular and systematically collected ophthalmic phenotypes at population scale.

To this end, we conducted a proteome-wide analysis of 2,920 circulating plasma proteins in 53,016 participants from the UK Biobank Pharma Proteomics Project. We assessed associations with 80 clinically defined ocular diseases and 51 quantitative eye traits, using multivariable-adjusted models to control for key covariates. We further evaluated the incremental predictive value of protein-based risk models and performed Mendelian randomization to infer causal relationships and prioritize therapeutic targets. Together, these analyses provide a comprehensive atlas of protein-ocular associations and aim to redefine the role of systemic biomarkers in the early detection, risk stratification, and mechanistic understanding of ocular diseases.

## Result

### Baseline characteristics

Among the 53,016 UKB-PPP participants (mean age: 56.8 years, 46.1% male, 93.4% of European ancestry; Table S1), a total of 2920 plasma proteins passed quality control for subsequent analysis (Table S2). Ocular diseases were classified into five anatomical categories: adnexal disorders, anterior segment disorders, glaucoma, posterior segment disorders, and refraction and other ocular disorders. The most prevalent diseases within each category included ptosis of eyelid (13.7%), cataract (unspecified, 34.6%), glaucoma (unspecified, 48.5%), degeneration of macula and posterior pole (32.2%), and astigmatism (42.3%) (Fig. 1A and Table S3).

**Fig. 1:**
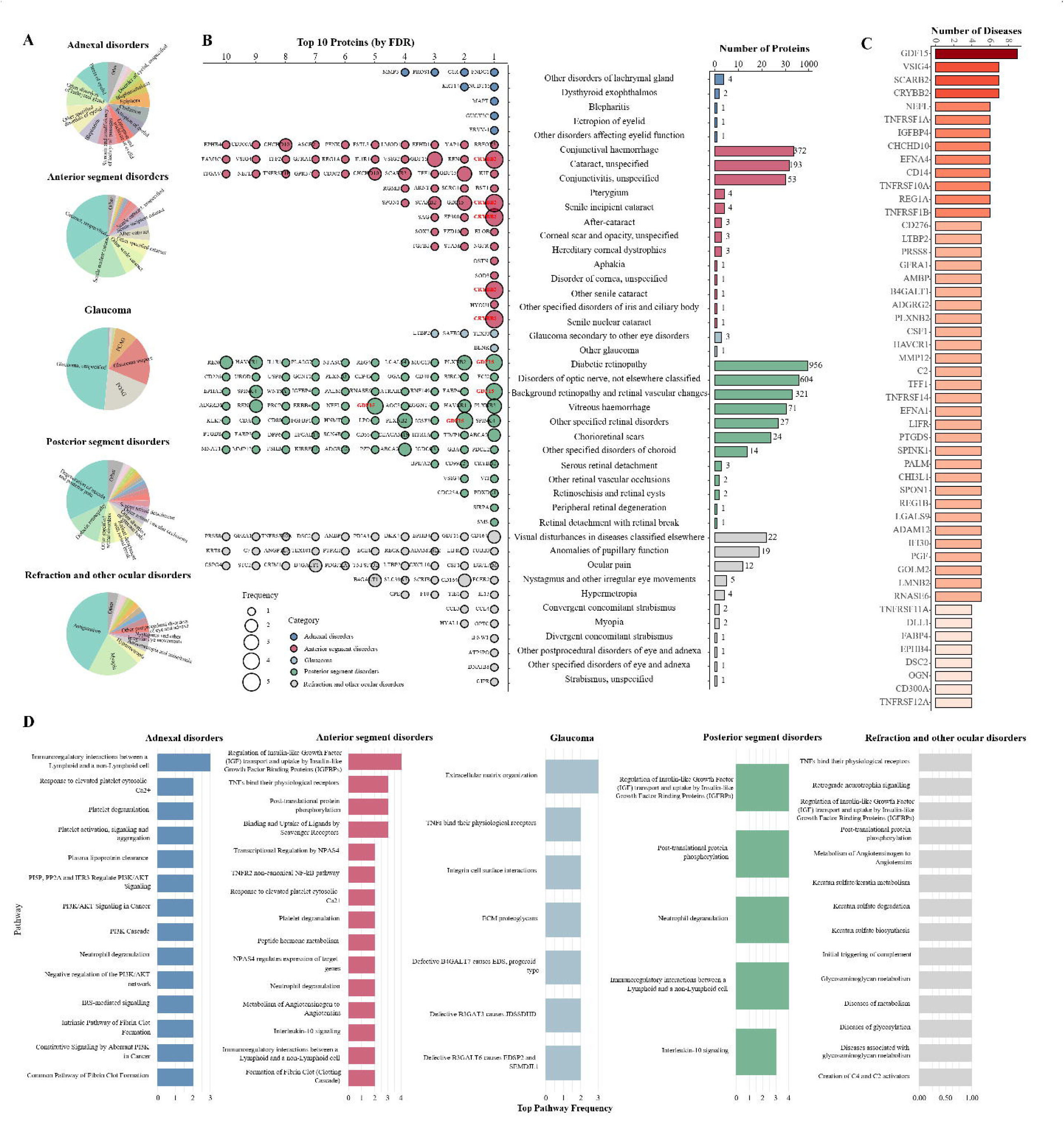
Summary of protein-disease associations across ocular diseases. **Note: A.** Ocular diseases were grouped into five major anatomical categories. Each pie chart displays the composition and relative proportion of specific diagnoses within each category. PCAG, primary angle-closure glaucoma; POAG, primary open-angle glaucoma. “Other” indicates diagnoses within each category with ≤20 positive cases, which were grouped together. **B.** The bar plot (right) shows the number of significantly associated proteins (FDR-adjusted P-value < 0.05) for each ocular disease. The dot plot (left) shows the top 10 proteins ranked by FDR per disease; if fewer than 10 proteins were significant, all are shown. Dot size represents the number of times a given protein appears across diseases within the same category, reflecting intra-category molecular sharing. **C.** Proteins are ranked by the number of ocular diseases with which they are significantly associated (FDR-adjusted P-value < 0.05). Only the top 40 proteins are displayed, highlighting those with the broadest cross-disease relevance. **D.** For each individual disease, Reactome pathway enrichment was conducted using significantly associated proteins (top 30 by FDR-adjusted P-value; if more than 30 proteins were significant, all were included). The top 10 enriched pathways per disease were retained, and pathway frequencies were aggregated within each disease category to identify shared or category-specific biological processes.

### Association of circulating proteins with ocular diseases

To ensure statistical robustness, ocular diseases with ≤20 cases were excluded, resulting in a final set of 80 conditions included in the primary analysis. Cox regression analyses adjusted for age and gender identified 2749 significant protein-disease associations across 80 ocular diseases after FDR correction (Fig. 1B and Table S4). The majority of significant associations occurred within anterior and posterior segment disorders, with diabetic retinopathy exhibiting the largest number (n=956).

Within anterior segment disorders, CRYBB2 demonstrated the strongest association cataract subtypes, consistent with previous evidence implicating CRYBB2 in the pathogenesis of multiple cataract types^16^ ^17^. In posterior segment disorders, growth differentiation factor 15 (GDF15), a member of the transforming growth factor-β (TGF-β) superfamily involved in insulin secretion, inflammation, and vascular homeostasis^18–21^, emerged as the most frequently associated protein. Recent studies suggest GDF15 expression during optic nerve injury and retinal neovascularization is regulated by hypoxia-inducible factors (HIFs), highlighting its relevance to diabetic retinopathy and retinal vascular pathology^21^ ^22^. Additionally, VEGFA and PGF, although not among the top 10 proteins associated with diabetic retinopathy, still exhibited significant associations (FDR-adjusted P=0.004 and P=5.479×10^-6^, respectively), further validating the robustness of our findings.

We next investigated the extent of protein sharing across ocular disease categories (Fig. 1C and Table S5). Several key proteins, notably GDF15, VSIG4, and SCARB2, emerged repeatedly across multiple disease categories, suggesting their roles as common biomarkers or therapeutic targets. In contrast, adnexal disorders, glaucoma, and refractive and other ocular disorders demonstrated limited protein overlap, indicating distinct underlying pathological processes. Pathway enrichment analysis identified disease category-specific biological pathways (Fig. 1D): anterior and posterior segment disorders primarily involved inflammatory, immune-regulatory, and vascular pathways, whereas glaucoma, adnexal disorders, and refractive and other ocular disorders showed enrichment in extracellular matrix remodeling, coagulation cascades, and neurotrophic factor signaling pathways.

To further dissect molecular similarities at the disease level, we constructed a pairwise protein overlap matrix across all ocular conditions and applied hierarchical clustering (Fig. S2). Anterior and posterior segment disorders exhibited substantial protein overlap, indicating shared molecular pathophysiology. In contrast, glaucoma and adnexal disorders showed minimal overlap. Refractive and other ocular disorders showed low overall overlap but were enriched for neurotrophin signaling, suggesting possible involvement of visual neural mechanisms.

### Association of circulating proteins with ocular health traits

We systematically evaluated the associations between 2920 plasma proteins and 51 ocular health traits (Table S7), identifying 3169 significant protein-trait pairs after FDR correction (Table S8). We first focused on three clinically relevant parameters: logMAR visual acuity, Intra ocular pressure by Goldmann-correlated (IOP_G), and intra ocular pressure by corneal-compensated (IOP_C). Each parameter showed significant associations with multiple plasma proteins, supporting a molecular link between ocular phenotypes and systemic proteomic signatures (Fig. 2A). A Venn diagram further showed substantial overlap between IOP_G and IOP_C, but minimal overlap with logMAR, indicating distinct molecular underpinnings of visual function and IOP regulation (Fig. 2B).

**Fig. 2:**
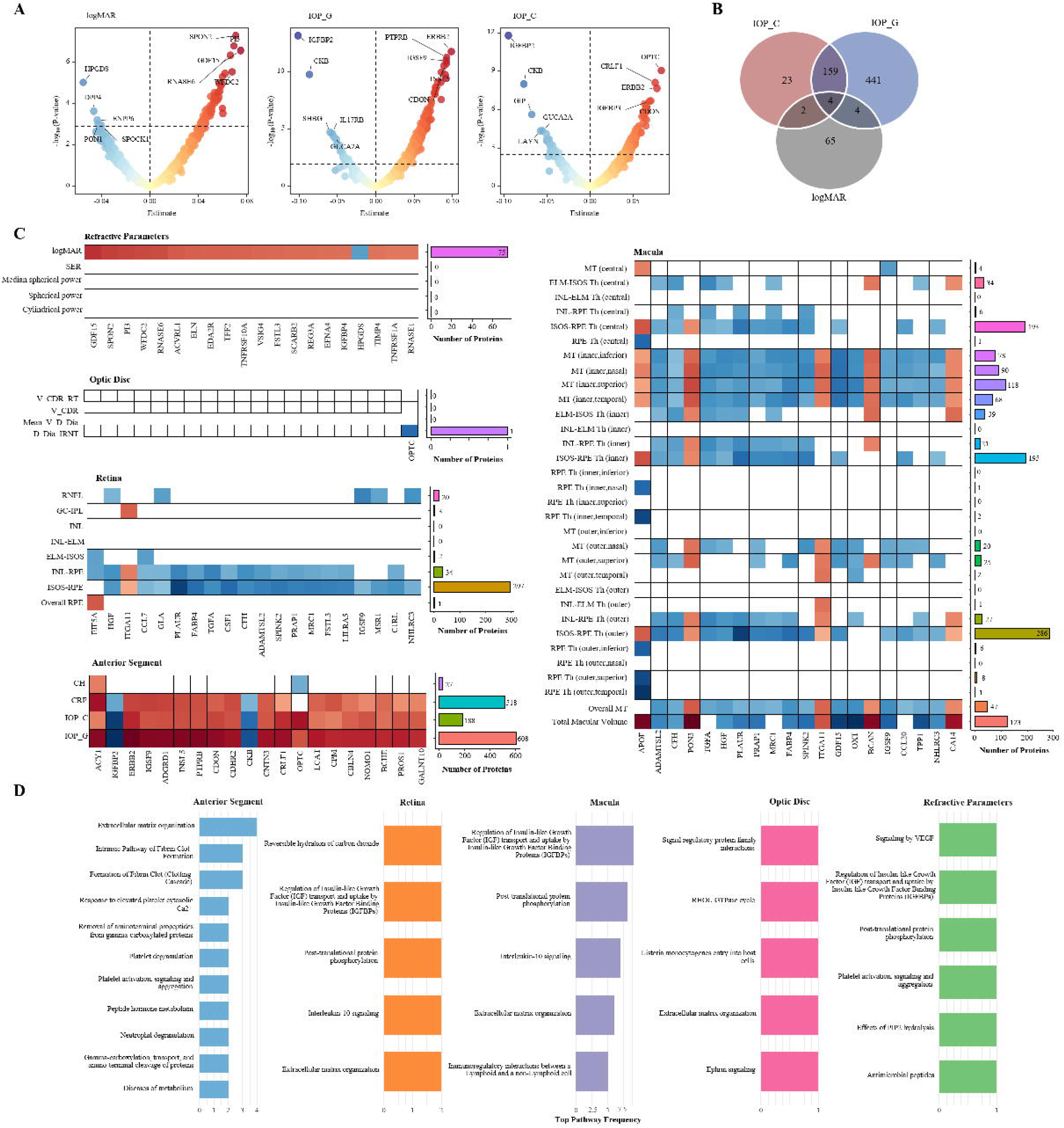
Summary of protein–trait associations across ocular health parameters. **Note: A.** Generalized linear models (GLMs) were used to assess associations between plasma proteins and three clinically relevant ocular traits: logMAR, IOP_G, and IOP_C. Volcano plots display the distribution of significantly associated proteins after FDR correction. IOP_G, Intra ocular pressure, Goldmann-correlated; IOP_C, Intra ocular pressure, corneal-compensated. **B.** A Venn diagram shows the overlap of significant proteins among the three traits. Substantial overlap was observed between IOP_G and IOP_C, while both shared fewer proteins with logMAR, suggesting distinct underlying molecular mechanisms. **C.** Ocular traits were grouped into five anatomical categories: refractive parameters, optic disc, retina, anterior segment, and macula. For each group, the number of significantly associated proteins was summarized (bar plot), and the top 20 proteins (ranked by the number of associated traits and absolute estimate size) were visualized in a heatmap to highlight protein–trait association patterns. **D.** Reactome pathway enrichment was performed for each trait using significantly associated proteins (top 30 by FDR-adjusted P value, or all if more than 30 were significant). The top 10 pathways per trait were retained, and pathway frequencies were aggregated within each anatomical category to identify shared or category-specific biological processes. Full ocular health trait names and abbreviations are listed in Supplementary Table S7.

To contextualize these associations, traits were grouped into five anatomical domains: refractive parameters, optic disc, retina, macula, and anterior segment (Fig. 2C). The anterior segment and macula exhibited the highest number of associated proteins, particularly for traits related to the macular inner ring and retinal ISOS-RPE thickness, highlighting these metabolically active regions as hotspots of systemic proteomic signals. In contrast, optic disc traits showed sparse associations, suggesting limited systemic involvement and greater local regulation.

In the top protein analysis (Fig. 2C), IGFBP2 and ERBB2 were the most prominent proteins associated with anterior segment traits. IGFBP2 showed a significant negative association with IOP_G (β=-0.102, P=1.946×10^-10^), potentially modulating intraocular pressure via IGF-1 regulation, while ERBB2 may influence trabecular meshwork remodeling through CD44H interactions^23^ ^24^. APOF was specifically associated with RPE thickness. As a plasma protein known to regulate cholesteryl ester transfer protein activity, APOF may play a role in lipid metabolism and contribute to retinal structural integrity^25^. PLAUR was negatively associated with ISOS-RPE thickness. As a central effector of the uPAR pathway, it has been linked to blood-retinal barrier (BRB) breakdown in diabetes^26^. Given that ISOS-RPE thickness serves as a proxy for photoreceptor outer segment integrity, which correlates with retinal function in diabetic macular edema, the association suggests a role for PLAUR in BRB disruption and photoreceptor degeneration^27^ ^28^.

Pathway enrichment analysis (Fig. 2D) revealed that retina- and macula-related traits were primarily enriched in growth factor signaling, immune responses, and extracellular matrix remodeling, while refractive and anterior segment traits were linked to structural and barrier-related pathways. In contrast, optic disc and refractive traits showed fewer protein associations and highly specific, non-overlapping pathways.

To further explore molecular relationships among traits, we constructed a protein overlap matrix and applied hierarchical clustering (Fig. S3, Table S9). Optic disc traits exhibited the greatest protein-level specificity with minimal sharing, while IOP and corneal biomechanical traits (e.g., CRF) shared many proteins, reflecting their physiological interdependence.

### The landscape of pleiotropy in protein-disease-trait association

To further investigate potential molecular hubs linking ocular health traits with disease outcomes, we constructed a plasma proteome-based pleiotropy map to systematically assess shared protein associations between eye diseases and ocular health parameters. We first focused on the observed relationship between ISOS-RPE thickness and diabetic retinopathy, which revealed 219 overlapping plasma proteins significantly associated with both phenotypes. Notably, the majority of these proteins exhibited effect directions consistent with ISOS-RPE thickness thinning and increased disease risk (Fig. 3A, 3B). For instance, elevated PLAUR levels were significantly associated with reduced ISOS-RPE thickness (β=-0.094, P=4.13×10^-8^) and heightened risk of DR (HR=1.367, P=1.81×10^-11^). Another protein of interest, PRAP1—relatively uncharacterized in ophthalmic research—demonstrated similar directional associations (β=-0.068, P=6.75×10^-5^; HR=2.122, P=1.42×10^-38^). As a newly described lipid-binding protein, PRAP1 has been shown to enhance MTTP-mediated lipid transport, and given the role of MTTP in retinal lipid metabolism, these findings suggest that PRAP1 may serve as a molecular link between retinal structural alterations and disease pathogenesis^29^ ^30^.

**Fig. 3:**
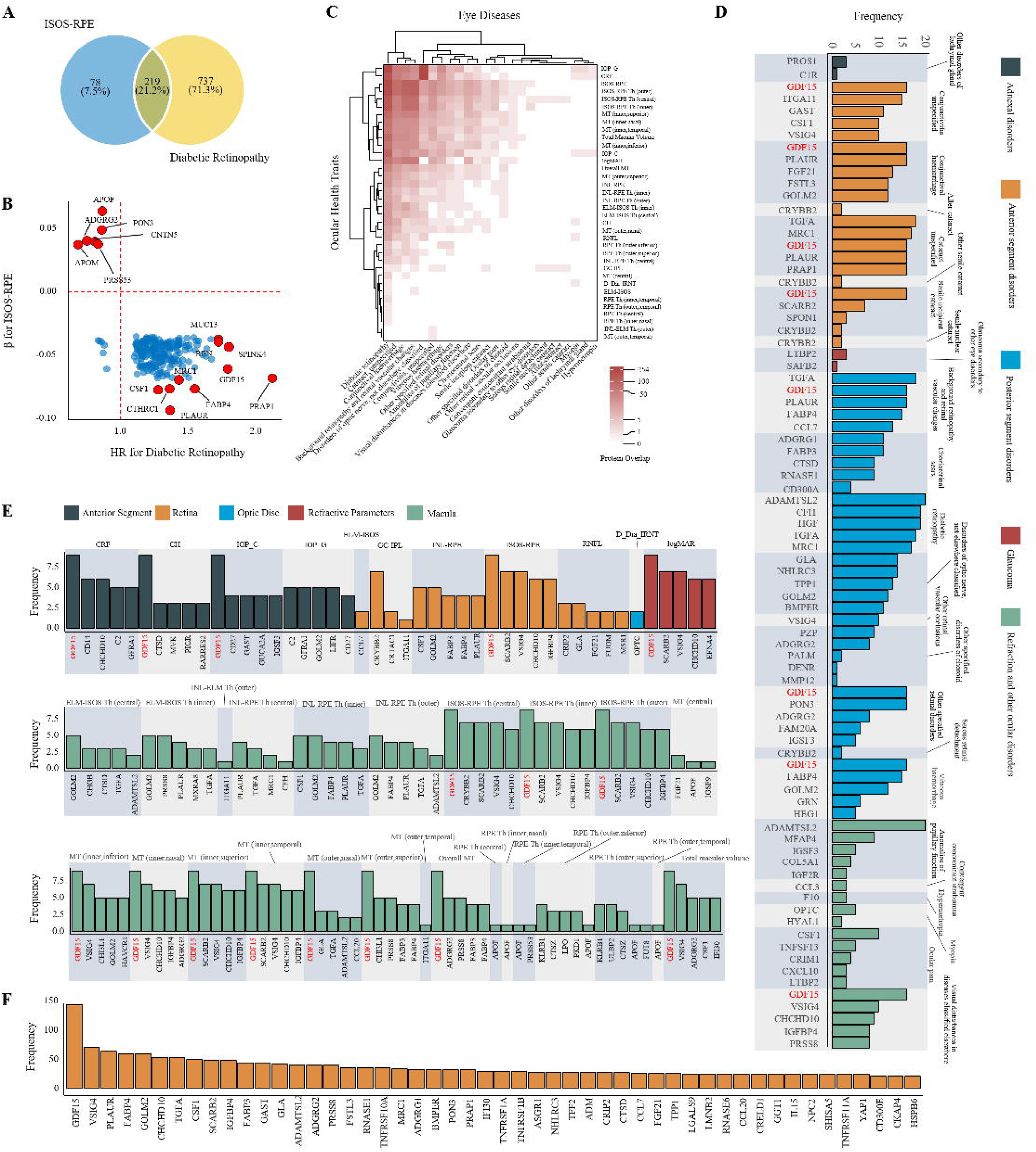
A proteomic atlas of pleiotropy across ocular diseases and health traits. **Note: A.** Venn diagram showing the overlap of significantly associated plasma proteins between ISOS-RPE thickness and diabetic retinopathy. **B.** Scatter plot displaying shared proteins between ISOS–RPE and diabetic retinopathy. Proteins with top five absolute β values (for ISOS-RPE) and the top five highest and lowest hazard ratios (for diabetic retinopathy) are highlighted. **C.** Heatmap of the number of shared proteins between ocular diseases and quantitative ocular health traits, clustered by pairwise protein overlap. **D.** Protein-level pleiotropy from a disease-centered perspective. For each disease, the top five most frequently shared proteins with ocular health traits are shown (if more than five had frequency > 5). **E.** Trait-centered view of pleiotropic proteins. For each ocular health trait, the top five most frequently shared proteins with ocular diseases are shown (if more than five had frequency > 5). **F.** Overall frequency of proteins shared between at least one ocular disease and one quantitative ocular trait.

Hierarchical clustering of shared protein associations between ocular diseases and health traits revealed that DR clustered closely with intraocular pressure, corneal biomechanical metrics, ISOS-RPE thickness, and other macular traits at the proteomic level (Fig. 3C, Table S10), suggesting that structural alterations may contribute to its systemic pathophysiology. From the disease perspective (Fig. 3D), anterior segment, posterior segment, and certain refractive disorders demonstrated frequent protein overlap with multiple ocular parameters. Notably, conjunctivitis and conjunctival haemorrhage shared a considerable number of proteins with macular structural parameters. Many of these frequently shared proteins—such as GDF15, VSIG4, PLAUR, FGF21, and FSTL3—are involved in immune regulation and inflammatory signaling pathways. Although direct mechanistic evidence is limited, animal studies have shown that conjunctival inflammation can induce retinal expression of inflammatory mediators such as TNF-α and IL-6^31^. These observations underscore the need to further explore immune-mediated crosstalk between anatomically distinct compartments of the eye. From the parameter perspective (Fig. 3E), macular traits—particularly ISOS-RPE thickness and inner ring thickness—showed the greatest degree of protein sharing across diseases, suggesting this region may represent a common structural vulnerability across ocular conditions. Among all proteins, GDF15 exhibited the strongest pleiotropy, being significantly associated with 144 distinct diseases-traits pairs (Fig. 3F). In view of its established role in systemic metabolic and inflammatory regulation, this finding positions GDF15 as a potential molecular bridge linking systemic homeostasis and ocular pathology^18^ ^21^.

### Protein-based prediction of ocular diseases

We evaluated the predictive performance of three models—based on proteome-only, clinical-only, and combined features—across 45 ocular diseases (Fig. 4, Table S11). Using the plasma proteomic model, seven diseases (15.6%) achieved an area under the curve (AUC) greater than 0.80, predominantly within anterior (3/13) and posterior (3/11) segment disorders. Diabetic retinopathy showed the highest discriminative performance with an AUC of 0.913 (95% CI: 0.878–0.938) (Fig. 4A). Compared to the clinical-only model, the proteome-only model yielded an AUC improvement of ≥0.02 in 20 diseases (44.4%), indicating enhanced predictive value. Adding proteomic features to the clinical model improved prediction in 23 diseases (51.1%), with the largest gains observed in posterior segment disorders (Fig. 4B). Notably, for most diseases, the combined model performed similarly to the proteome-only models. Although four diseases showed marginally higher AUCs in the combined model, the maximum difference was only 0.021, suggesting that plasma proteomics accounts for the majority of predictive power.

**Fig. 4:**
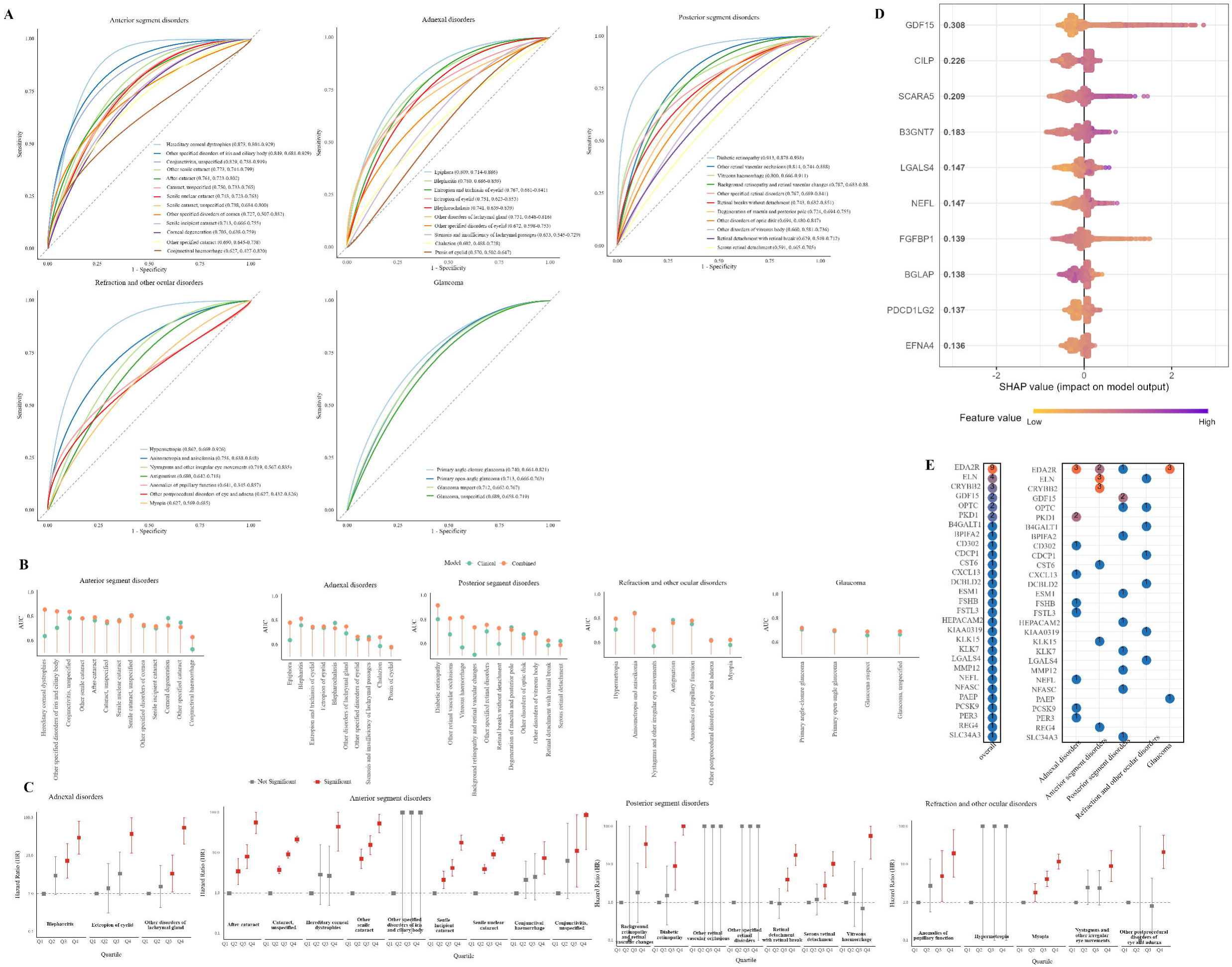
Evaluation of proteome-based prediction of ocular diseases and its added value beyond clinical risk factors. **Note: A.** Receiver operating characteristic (ROC) curves for disease prediction across different anatomical categories using models trained on the full plasma proteome. Values in parentheses indicate the area under the curve (AUC) with 95% confidence intervals (CI). **B.** Comparison of AUCs for models using clinical risk factors alone (green) versus combined models incorporating proteomic features (orange). **C.** Based on the proteome-only prediction model, ocular diseases with at least one significantly associated protein were selected for further analysis. Participants were stratified into quartiles (Q1-Q4) according to the proteomic risk score, and hazard ratios (HRs) were estimated using Cox proportional hazards models. Q1 served as the reference group (HR = 1, shown in gray); red squares in Q2-Q4 indicate significantly elevated risk compared to Q1 (P□<□0.05), while gray squares indicate non-significant associations. **D.** SHAP values were used to rank the most influential proteins in the predictive model for diabetic retinopathy. The x-axis represents the contribution of each protein to the model output. **E.** Proteins were prioritized based on SHAP-derived importance across all diseases. The left panel summarizes the number of times each protein ranked first in SHAP value across all prediction models. The right panel shows the distribution of these top 29 proteins across different ocular disease categories, with dot color representing their frequency of occurrence within each category.

We further assessed the relationship between proteomic risk scores and the incidence of ocular diseases (Fig. 4C). Among diseases with at least one significantly associated protein, several showed markedly higher incidence rates in the highest quartile (Q4) of proteomic scores compared to the lowest (Q1) (PL<L0.05), supporting the potential utility of proteomic profiling for population-level risk stratification and early identification of high-risk individuals. Notably, in multiple cataract-related conditions (e.g., unspecified cataract and senile nuclear cataract), the proteome-based prediction models yielded only moderate AUCs; however, the corresponding risk scores demonstrated clear dose-response relationships with disease incidence. This suggests that proteomic signals may sensitively capture subclinical or progressive molecular changes not fully reflected in discriminative performance. In contrast, for certain diseases such as hypermetropia, no significant gradient was observed, potentially due to dispersed score distributions or non-linear pathophysiological associations. Importantly, for diabetic retinopathy—which achieved the highest AUC among all outcomes—we observed both strong discriminative power and a consistent dose–response pattern, underscoring the robustness and clinical relevance of proteomic risk signatures for this condition.

We assessed the relative importance of each protein in the disease prediction models (Table S11). For diabetic retinopathy, SHAP analysis identified GDF15 as the top contributing feature (Fig. 4D). This protein was not only significantly associated in univariate analyses but also ranked highest in the predictive model, highlighting its strong and consistent predictive value for this condition. Across all models, EDA2R emerged most frequently as the top SHAP-ranked protein, appearing in nine diseases, followed by ELN, CRYBB2, GDF15, and OPTC (Fig. 4E). Although GDF15 showed the broadest range of associations, it ranked first in SHAP less frequently and primarily in posterior segment diseases, suggesting a more general but less disease-specific predictive role. In contrast, EDA2R was associated with fewer diseases in univariate analyses but demonstrated strong model-based importance, indicating a potentially central regulatory role in certain pathological pathways, rather than functioning as a weak, broadly linked marker. Notably, EDA2R was not significantly associated with any glaucoma-related phenotype in univariate testing, yet was ranked as the top predictor in its SHAP profile. Prior studies have shown that the Ectodysplasin A signaling pathway contributes to ocular surface homeostasis, with EDA2R acting as its key receptor, offering biological plausibility for its potential role in ocular pathophysiology^32^.

### Hierarchical clustering reveals systemic modules across eye diseases and traits

To resolve shared molecular patterns across clinically defined eye diseases and quantitative ocular parameters, we applied hierarchical clustering to scaled protein-trait associations across 43 diseases and 37 ocular traits. This analysis identified six proteome-defined ocular modules (Fig. 5A), each comprising anatomically diverse yet molecularly convergent phenotypes. The optimal number of clusters was determined based on dendrogram topology, silhouette width, and gap statistic (Fig. S4-S5).

**Fig. 5:**
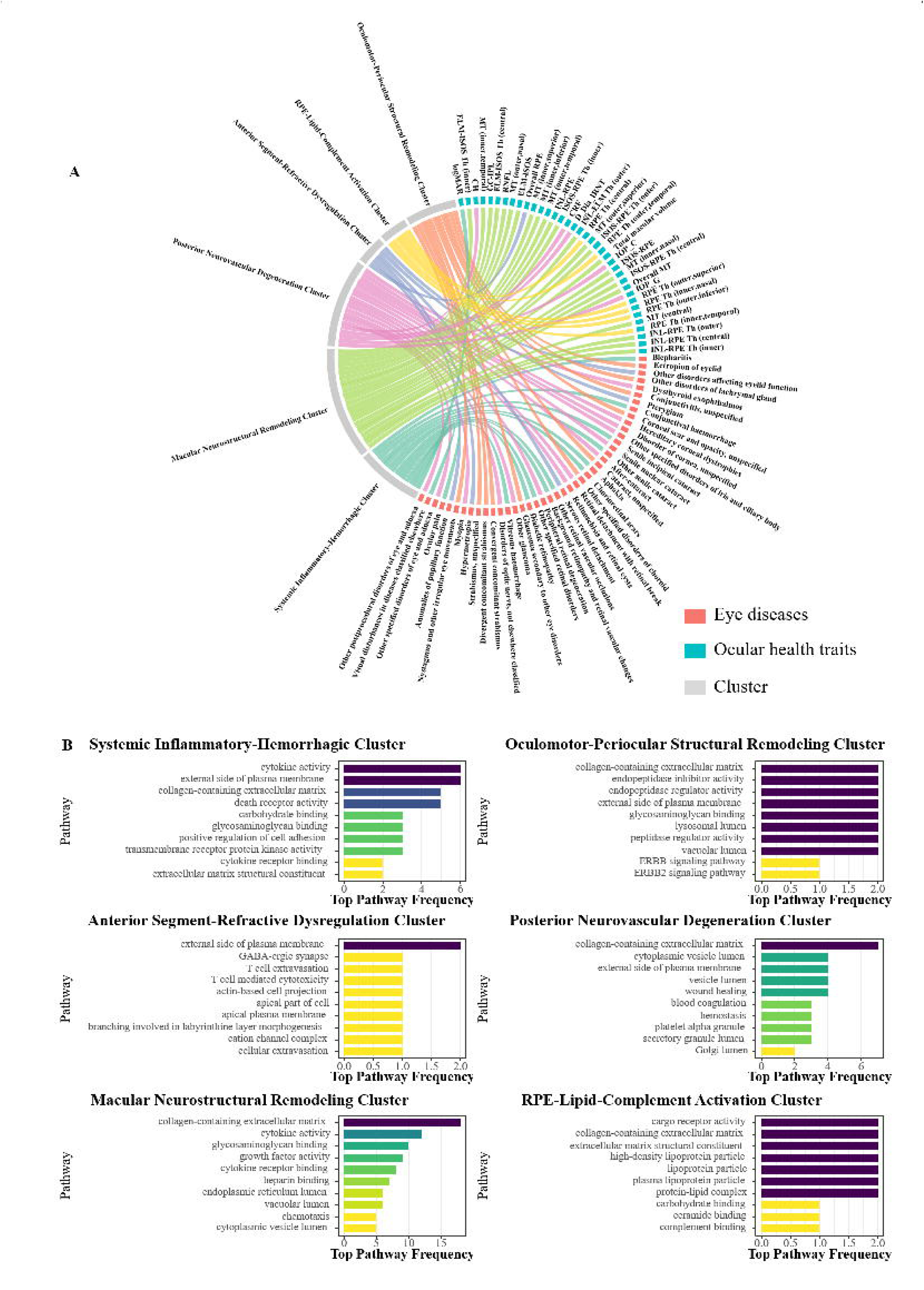
Clustered ocular phenotypes reveal shared proteomic and biological signatures. **Note: A.** Circular plot showing the clustering of 43 eye diseases (blue) and 37 ocular traits (red) based on protein association profiles, identifying six distinct clusters (colors). Links represent shared proteins between phenotypes and clusters. B. Top enriched GO biological processes and cellular components within each cluster, based on the frequency of appearance across cluster-associated proteins. Each bar indicates the top pathways enriched per cluster, with stronger enrichment denoted by higher frequency counts.

The Systemic Inflammatory-Hemorrhagic Cluster grouped diabetic retinopathy, vitreous haemorrhage, and visual loss, enriched in cytokine signaling, ECM remodeling, and death receptor pathways—pointing to a systemic immune-vascular program driving retinal injury. The Oculomotor-Periocular Structural Remodeling Cluster included strabismus, nystagmus, eyelid malposition, and periocular inflammation, with enrichment in ERBB and peptidase pathways, implicating disrupted extracellular architecture and motor coordination. The Anterior Segment-Refractive Dysregulation Cluster aggregated hypermetropia, pterygium, pupillary anomalies, and RPE thickening, linked to synaptic, cytotoxic, and epithelial polarity pathways—suggesting surface instability at a neuroimmune-epithelial interface. The Macular Neurostructural Remodeling Cluster, captured inner retinal and macular thickness metrics (e.g., GC-IPL, ISOS-RPE), enriched in collagen assembly, cytokine binding, and ER stress, reflecting neurostructural remodeling under chronic stress. The Posterior Neurovascular Degeneration Cluster encompassed glaucoma, optic neuropathies, senile cataract, corneal and periocular disorders, and biomechanical traits (IOP, CH, CRF), unified by vesicle transport, coagulation, and platelet activation pathways—indicating a convergence of structural fragility, mechanical stress, and neurovascular compromise. Finally, the RPE-Lipid-Complement Activation Cluster captured regional RPE thickness variations, enriched in complement activation, lipoprotein metabolism, and cargo transport, highlighting an RPE-centered lipid–immune axis relevant to macular pathology.

Together, this unsupervised clustering defines a modular proteomic taxonomy of the ocular phenome, linking clinically and anatomically distinct traits to shared systemic biology. The pathway coherence across clusters reinforces the relevance of circulating proteins in organizing ocular phenotypes into actionable, biologically meaningful subgroups for biomarker discovery and therapeutic targeting.

### Potential causal proteins of eye diseases and ocular health traits

To further elucidate the potential pathogenic mechanisms linking ocular diseases and physiological traits, we conducted Mendelian randomization (MR) analyses using cis-acting pQTLs as instrumental variables, based on pQTL data and GWAS summary statistics. The analysis focused on previously identified significant protein-trait associations (including both diseases and quantitative parameters). A few outcomes with insufficient GWAS data or too few associated proteins were excluded (Table S12). Overall, we identified 144 protein-disease pairs and 122 protein–trait pairs showing potential causal relationships (P < 0.05). Diabetic retinopathy (n = 62 significant proteins), disorders of optic nerve, not elsewhere classified (n = 32), CRF (n = 43), and ISOS-RPE thickness-related parameters (n = 49) exhibited relatively high numbers of causal proteins (Fig. 6A-B; Tables S13-S14), providing additional causal evidence for the protein-outcome associations identified in previous analyses.

**Fig 6.**
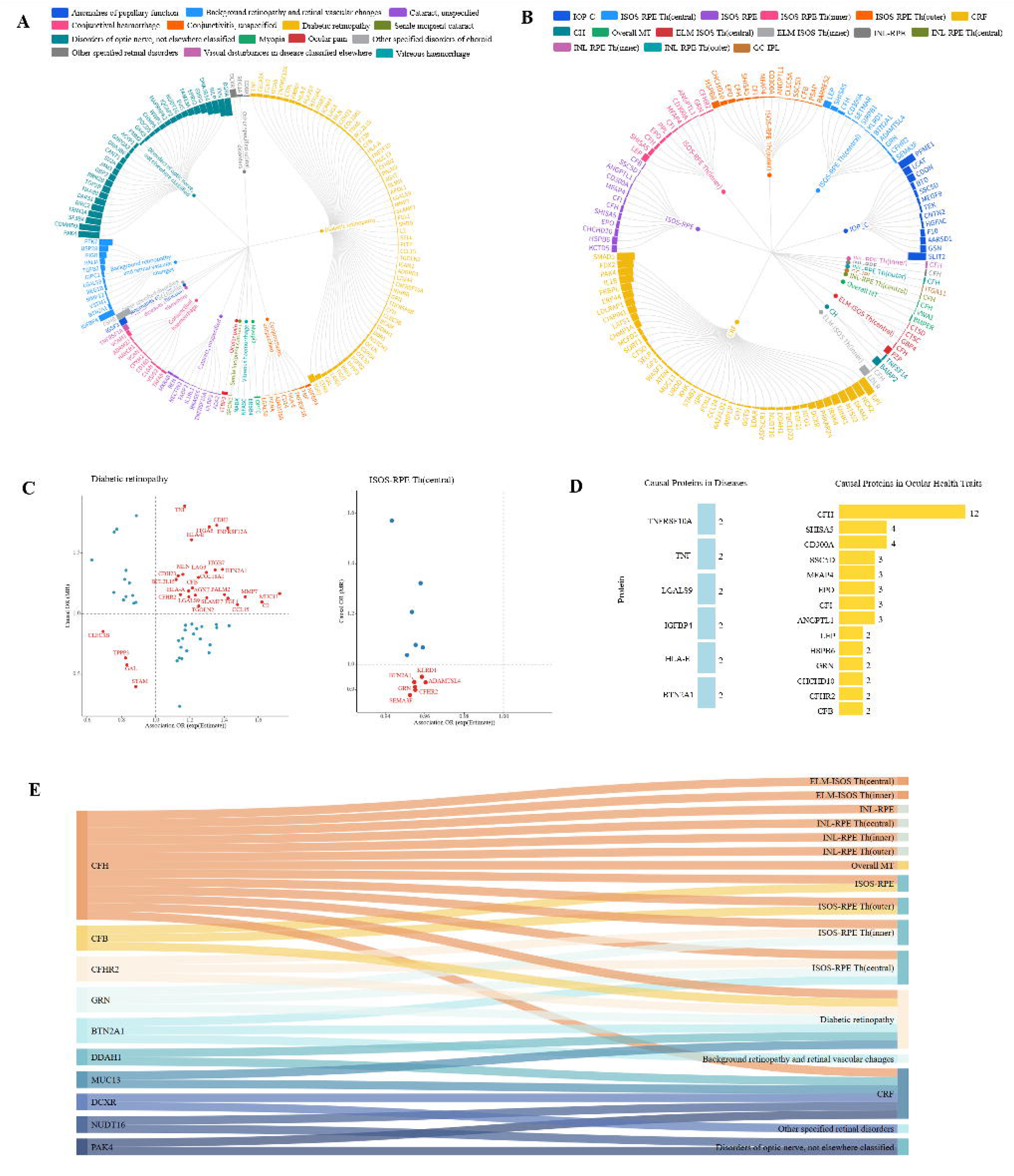
Summary of potential causal proteins for eye diseases and ocular health traits through cis-MR analysis. **Notes: A-B.** Radial tree plots show significant protein-disease pairs **(A)** and protein-ocular health trait pairs **(B)** identified through cis-MR, with all pairs also reaching significance in association analyses. Each bar represents the direction and magnitude of the MR effect estimate; outward bars indicate positive effects and inward bars indicate negative effects. Outer rings display the significant proteins, and inner rings show the corresponding diseases **(A)** or ocular health traits **(B)**, colored by phenotype category. **C.** Scatter plots depict the consistency of effect direction between association and MR results. Red dots indicate proteins with concordant directions (i.e., both ORs >1 or <1). Shown for two representative traits: diabetic retinopathy and ISOS-RPE thickness (central). **D.** Bar plots summarize the frequency of proteins identified as causal across multiple phenotypes. The left panel shows diseases, and the right panel shows ocular health traits, suggesting potential pleiotropic roles. **E.** Sankey plot illustrates the overlap of causal proteins with significant results in both association and MR analyses, across diseases and ocular health traits. Left: protein names; right: associated phenotypes.

Among all protein-trait causal associations, we highlight two representative phenotypes with the most causal proteins: diabetic retinopathy and central ISOS-RPE thickness (Fig. 6C). In diabetic retinopathy, 29 proteins (46.8%) showed consistent effect directions between MR and effect estimates from association analyses; for ISOS-RPE Th(central), 6 proteins (50.0%) showed such concordance. Several proteins, such as TNFRSF12A^33^, CDH2^34^, and TNF^35^, have been previously implicated in the pathogenesis of diabetic retinopathy, while SEMA3F may exert protective effects on macular integrity by inhibiting outer retinal/choroidal neovascularization^36^. Causal frequency analysis further revealed that CFH was repeatedly identified as causal across 12 ocular parameters—mostly related to outer retinal structure—suggesting a core role in maintaining retinal architecture. Previous studies have shown that CFH deficiency disrupts organelle distribution in retinal pigment epithelium and alters rod photoreceptor outer segment morphology^37^. In contrast, causal proteins involved in ocular diseases showed limited overlap, with a maximum of two diseases sharing the same protein, indicating a higher phenotype-specificity in disease-related causal pathways (Fig. 6D). Trait-overlapping analyses revealed that CFH, CFB, and CFHR2 were repeatedly implicated in both ISOS–RPE thickness and diabetic retinopathy, pointing to complement regulation as a potential shared mechanism linking structural integrity and disease pathogenesis. Additionally, CRF shared several causal proteins with both diabetic retinopathy and optic nerve disorders (e.g., DCXR, NUDT16, and PAK4), suggesting that corneal biomechanical properties may influence posterior segment structure through shared biological pathways, underscoring the importance of mechanical homeostasis in ocular health (Fig. 6E).

## Discussion

This study is the first to systematically map the complex and selective molecular associations between plasma proteins and ocular phenotypes, spanning both ocular diseases and structural traits, in 53,016 individuals. We identified 2,749 protein-disease associations, predominantly involving vascular disorders of the anterior and posterior segments, with diabetic retinopathy being the most prominent. Among 51 ocular health traits, 3,169 protein-trait associations were found, particularly for ISOS-RPE thickness, CRF, and IOP, highlighting a tight connection between systemic proteins and intraocular structural integrity. Further analyses revealed substantial protein overlap between vascular eye diseases such as DR and structural traits such as ISOS-RPE thickness and CRF, with some proteins showing potential causal effects. However, proteins significant across association, prediction, and causality analyses overlapped only minimally, suggesting functional heterogeneity. For example, GDF15 was broadly associated with diverse traits, EDA2R was identified as a key predictive feature despite weak marginal associations, and CFH consistently showed causal effects across retinal structural traits. These findings provide the most comprehensive atlas to date linking the plasma proteome to ocular phenotypes and uncover molecular pathways that may inform precision diagnostics and targeted interventions.

Traditionally, the blood-ocular barrier has been considered a major obstacle preventing large plasma proteins from accessing intraocular compartments, limiting their use as biomarkers for eye health. However, Freddo et al^38^. reported that aqueous humor secretion and plasma protein entry are “semi-independent events,” with approximately 1 percent of plasma proteins able to diffuse through ciliary capillaries into the iris stroma under physiological conditions, where posterior iris tight junctions and aqueous outflow prevent further penetration into the posterior chamber. A recent multi-omics study published in Cell identified 1,920 ocular cell-type marker proteins in the vitreous and aqueous humor, including 128 proteins expressed exclusively in hepatocytes, providing strong evidence for plasma proteins crossing ocular barriers^39^. Under pathological conditions such as diabetic retinopathy, the blood–retinal barrier (BRB) becomes disrupted due to endothelial and pericyte dysfunction and elevated inflammatory mediators, allowing plasma protein leakage into the retina and contributing to hard exudates^40–43^. Our findings further support this, revealing widespread associations between plasma proteins and both anterior and posterior segment structural parameters, including CRF, IOP, and ISOS-RPE thickness. These associations were particularly enriched in diseases affecting anterior and posterior segments, such as conjunctival hemorrhage and DR. In addition, posterior segment pathways related to inflammation, extracellular matrix remodeling, and cell adhesion were significantly enriched, consistent with early BRB disruption mechanisms in retinal vascular diseases. Together, these observations challenge the notion that plasma proteins are uninformative for ocular conditions and suggest their potential as non-invasive, scalable biomarkers for eye disease.

In this study, we identified substantial overlap in plasma proteins between anterior and posterior segment vascular diseases, particularly DR, and key structural parameters including ISOS-RPE thickness, CRF, and IOP. These findings suggest that circulating proteins may reflect systemic health and detect early microstructural degeneration within the eye, which is especially critical for early diagnosis. ISOS-RPE thickness, an indicator of photoreceptor outer segment and retinal pigment epithelium integrity, has received limited attention but has recently been shown to decrease in early-stage diabetic retinopathy, supporting its role as a structural marker of early microvascular dysfunction^44^ ^45^. Among shared proteins across phenotype s, GDF15, VSIG4, and PLAUR were prominent. While each has been individually implicated in inflammation regulation, immune homeostasis, or blood-retinal barrier function^21^ ^26^ ^46^, our study is the first to integrate them as common molecular signatures linking structure and disease. Notably, GDF15 has been reported to be elevated in the plasma of patients with diabetic retinopathy, with levels correlating with disease severity^47^. CRF, a biomechanical parameter reflecting corneal and intraocular pressure homeostasis, also shared proteins with diabetic retinopathy and several retinal traits, further supporting its potential as a systemic interface marker. These findings expand the utility of plasma proteins for precise ocular screening and provide a focused molecular basis for future mechanistic and interventional studies.

Although ISOS-RPE thickness exhibited substantial protein overlap with DR—including several proteins with inferred causal roles—the two traits did not cluster together in our hierarchical analysis. Rather than undermining their association, this suggests that ISOS-RPE and DR may represent anatomically distinct but molecularly convergent responses to shared systemic stressors such as chronic inflammation or metabolic dysfunction. In contrast, traits like IOP and CRF clustered tightly with other glaucoma, disorders of optic nerve, and senile nuclear cataract, pointing to a unified axis of biomechanical stress and neurodegeneration. The dissociation between ISOS-RPE and DR, despite their molecular overlap, underscores the potential of certain structural parameters to serve as early sentinels of systemic disease—reflecting upstream systemic influences rather than being classical downstream outcomes. More broadly, these findings suggest that molecular clustering of ocular traits is shaped more by shared systemic pathways than by anatomical contiguity, offering a framework for pan-disease biomarker discovery and precision screening.

Across multiple ocular diseases, the predictive potential of the plasma proteome was systematically evaluated, with particularly strong performance observed in diabetic retinopathy. Our analysis showed that plasma proteins not only effectively predicted the onset of DR but also provided added predictive value beyond traditional clinical variables for several anterior and posterior segment diseases, significantly enhancing risk stratification. Notably, even modest improvements in predictive performance may help detect early changes in ocular health. These findings highlight the potential of plasma proteins as accessible early warning markers, offering new avenues for proactive prevention and personalized intervention with clinical relevance.

Through MR analyses, we identified a set of plasma proteins with potential causal effects on ocular diseases and ocular health parameters. Most of these proteins are secreted or small-structured, suggesting good druggability. For example, CFH and CFB act as negative and positive regulators of the complement system, respectively, and several related therapies (e.g., Danicopan, Iptacopan, IONIS-FB-LRx) are currently in clinical trials, demonstrating therapeutic potential for diseases such as AMD^48–50^. DCXR, an intracellular carbon metabolism enzyme^51^, points to a potential link between detoxification of glycation byproducts and ocular disease pathogenesis. Although the blood-ocular barrier restricts the direct impact of large proteins, evidence suggests that under pathological conditions, mechanisms such as endothelial transcytosis, inflammation-induced barrier disruption, and neuro-immune signaling may enable systemic proteins to influence intraocular structures. These causal proteins not only reveal systemic regulatory mechanisms underlying ocular disease but also inform future therapeutic target development at the systemic level.

Among ocular disease categories labeled as “unspecified,” “other,” or “not elsewhere classified,” we observed many significant protein associations, suggesting that plasma proteins may capture pathological states that elude conventional diagnostic classifications. These nonspecific categories likely reflect the complexity and heterogeneity of ocular conditions, which are often difficult to define precisely using traditional clinical criteria. As systemic indicators^6–8^, plasma proteins may uncover links between ocular disease and broader metabolic or inflammatory processes. Therefore, profiling plasma protein patterns may support a new classification framework for eye diseases while also serving as a proxy for systemic health. This finding underscores the concept of the eye as a potential “sensor” of systemic disease and expands the scope of ophthalmic research toward a more integrated view of systemic and ocular health.

This study leveraged large-scale proteomic data to systematically map associations between plasma proteins and a broad range of ocular diseases and health parameters, spanning trait associations, cross-phenotype mechanisms, predictive performance, and causal inference—highlighting a strong systems medicine orientation. However, several limitations remain. First, although plasma proteins are accessible and clinically translatable, their indirect nature may limit their ability to fully reflect intraocular dynamics, particularly in highly localized anterior or posterior segment conditions. Second, MR analyses relied on cis-pQTLs as instruments, and some proteins were supported by weak or single-variant instruments, potentially reducing the robustness of causal estimates^52^. Third, predictive models were not validated in independent cohorts, limiting their generalizability. Lastly, the study population was predominantly of European ancestry, and the transferability of findings to other populations remains uncertain. Future studies should integrate intraocular fluid proteomics, single-cell and spatial transcriptomics to dissect system-local interactions, and employ prospective, multi-ethnic validation to advance the clinical translation of plasma proteins in early detection, subtyping, and targeted treatment of ocular diseases.

Leveraging large-scale population proteomic data, we delineated a comprehensive map of plasma protein associations with diverse ocular diseases and structural parameters. Key proteins with predictive relevance and genetic causal support were identified, forming an integrated framework of cross-phenotype mechanisms and potential therapeutic targets. Underrecognized ocular parameters, such as ISOS-RPE thickness and CRF, emerged as critical mediators linking systemic and local processes. Additionally, several “not elsewhere classified” eye conditions displayed distinct proteomic patterns, pointing to possible plasma protein–driven molecular subtypes. These insights strengthen the concept of the eye as a systemic health sensor and broaden the boundaries of ophthalmic research toward molecular diagnostics and mechanistic understanding. Moving forward, plasma proteomics may support early detection, refined classification, and targeted treatment of eye diseases, promoting convergence between ophthalmology and systems medicine.

## Conclusion

This study redefines the molecular architecture of ocular phenotypes by demonstrating that the plasma proteome reflects not only local eye pathology but also integrates systemic immunometabolic and biomechanical signals. Through large-scale profiling, we uncovered widespread and biologically coherent links between circulating proteins and diverse ocular traits—revisiting the traditional view that the blood-ocular barrier limits the relevance of systemic biomarkers. The identification of shared protein signatures, putative causal mediators, and modular phenotypic clusters suggests that certain eye diseases may be better conceptualized as systemic end-organ manifestations rather than isolated ocular conditions. These findings lay a molecular foundation for precision diagnostics, stratified screening, and therapeutic repurposing in ophthalmology, guided by systemic proteomic insights.

## Methods

### Study Design and Participants

This study leveraged data from the UK Biobank (UKB), a large-scale prospective cohort of ∼500,000 individuals recruited across the United Kingdom between 2006 and 2010. Plasma proteomic profiling was conducted on a randomized subset of participants through the UK Biobank Pharma Proteomics Project (UKB-PPP). Our analysis included 53,016 participants with available proteomic data and ophthalmic phenotypes, including clinical eye disease diagnoses classified according to the International Classification of Diseases, Tenth Revision (ICD-10 codes H00–H59) and ocular imaging parameters from optical coherence tomography (OCT). Fig S1 provides an overview of the study design and analytical workflow. Ethical approval for UKB was granted by the North West Multi-Centre Research Ethics Committee (REC reference: 11/NW/0382), and this study was conducted under UKB application number 515707.

### Data Collection and Preprocessing

#### Protein Measurements and Proteomic Data Processing

Proteomic profiling was conducted on EDTA-plasma samples from 54,893 participants in the UKB-PPP using the Olink Explore 1536 and Expansion platforms, quantifying 2,923 unique proteins. Following established protocols^53^, participants and proteins with more than 50% missing data were excluded, retaining 2,920 proteins and 53,016 participants for subsequent analyses. Missing data were imputed via chained random forests, utilizing age and sex as predictors. Protein expression levels, represented as Normalized Protein eXpression (NPX) values, were standardized to a mean of 0 and a standard deviation of 1.

#### Eye Diseases Ascertainment

Incident eye diseases were identified using ICD-10 diagnostic codes (H00–H59) from UK Biobank electronic health records, including hospital inpatient admissions and primary care consultations. Participants diagnosed with any eye disease at baseline or before UKB enrollment were excluded, resulting in disease-specific cohorts (summarized in Supplementary Table S1). Diagnoses were grouped into five clinically relevant categories: adnexal disorders, anterior segment disorders, glaucoma, posterior segment disorders, and refraction and other ocular disorders. Complete ICD-10 code mappings for these categories are detailed in Supplementary Table S3.

#### Ocular Parameters Ascertainment

OCT-derived eye parameters were obtained from values extracted by external researchers using UK Biobank (UKB) retinal optical coherence tomography (OCT) images captured with the Topcon 3D OCT-1000 device, as provided in the UKB Data Showcase and validated in prior studies^54^ ^55^ (see UKB returns catalogue for details). Parameters were classified into five categories based on anatomical and functional relevance to ocular structures: Anterior Segment (e.g., corneal hysteresis), Macula (e.g., macular thickness at the central subfield), Optic Disc (e.g., vertical cup to disc ratio), Refractive Parameters (e.g., logMAR), and Retina (e.g., average retinal nerve fiber layer thickness). This classification facilitates targeted analyses of distinct ocular regions implicated in various eye diseases. For parameters reported separately for left and right eyes (e.g., macular thickness at the central subfield, average retinal nerve fiber layer thickness), mean values were calculated for subsequent analyses. Due to partial overlap between samples with eye parameters and proteomic data, analyses were restricted to 53,016 participants with both datasets available. A full list of eye parameters (e.g., sample sizes per parameter detailed) is provided in Supplementary Table S7.

#### Covariates Ascertainment

Covariates included age (continuous), gender (male or female), ethnicity (self-reported), education (categorical: high, intermediate and low), smoking status (binary: never/previous or current), alcohol intake frequency (binary: not daily or daily/almost daily), sleep duration (binary: unnormal or normal), vegetable intake (binary: insufficient or sufficient), fruit intake (binary: insufficient or sufficient), regular physical activity (binary: yes or no), body mass index (BMI) (continuous), sleeplessness (binary: yes or no), social contact (binary: low or high), fed up feeling (binary: yes or no), tiredness (binary: yes or no), nerves (binary: yes or no), life stress (binary: yes or no) and townsend deprivation index (categorical: high, medium, or low).

### Analytical Methods

#### Proteomic Association Analyses

##### Proteomic-Eye Diseases Association Analyses

Cox proportional hazards regression models were used to assess associations between baseline plasma protein levels and the risk of incident eye diseases. Individuals with a diagnosis of the corresponding eye disease at baseline were excluded from each respective analysis. Each protein was tested separately, with time to first diagnosis as the outcome and follow-up spanning from baseline (2006–2010) to December 2024. All models were adjusted for the covariates described above, including ascertainment factors. Hazard ratios (HRs) and 95% confidence intervals (CIs) were estimated. Analyses were restricted to diseases with more than 20 incident cases. Statistical significance was determined using a false discovery rate (FDR)-corrected p-value threshold of <0.05.

##### Proteomic-Eye Parameters Association Analyses

Generalized linear models (GLMs) with Gaussian distributions were used to evaluate associations between plasma protein levels and OCT-derived ocular parameters. For each ocular parameter, only the first available measurement per participant was included in the analysis, regardless of assessment year. Each of the 2,920 proteins was tested independently against each ocular parameter as the outcome. Models were adjusted for all listed covariates, and beta coefficients with 95% confidence intervals (CIs) were estimated. Statistical significance was defined by an FDR-corrected p-value threshold of <0.05.

#### Shared Proteins across eye diseases and eye parameters

To explore molecular overlap, we identified and quantified shared proteins between each pair of eye diseases and ocular parameters. Sharing was defined by the number of proteins significantly associated with both traits. Cluster analyses were then performed based on the matrix of shared protein counts to uncover patterns of molecular similarity between diseases and parameters. Proteins shared across multiple diseases and parameters were ranked by frequency to highlight key cross-cutting molecules. Additionally, we conducted separate clustering analyses within the disease group and the ocular parameter group to characterize internal patterns of protein sharing. All analyses were based on the counts of shared significant proteins between traits.

#### Pathway Enrichment Analyses

We conducted pathway enrichment analyses using Gene Ontology (GO), KEGG, and Reactome databases for each eye disease and ocular parameter separately. Analyses were performed on the set of proteins significantly associated with each outcome. For outcomes with fewer than 30 significant proteins, the top 30 ranked by association p-values were included to ensure analytical consistency. Reactome enrichment was prioritized for downstream interpretation due to its detailed pathway granularity and disease relevance. For each outcome (disease or parameter), we identified the top 10 enriched Reactome pathways based on FDR-corrected significance. To highlight commonly dysregulated biological processes, we then summarized the most frequently recurring pathways across all outcomes. These analyses enabled the identification of shared and distinct biological mechanisms underlying different eye traits at the pathway level.

#### Mendelian Randomization

We implemented a two-sample Mendelian Randomization (MR) framework to investigate the causal relationships between plasma protein concentrations and a spectrum of ophthalmic disorders and ocular characteristics. Genetic instruments, specifically protein quantitative trait loci (pQTLs), were acquired from the UK Biobank Pharma Proteomics Project (UKB-PPP). For the outcomes, which encompassed eye-related diseases and quantitative ocular traits such as intraocular pressure and refractive error, we utilized summary statistics from genome-wide association studies (GWAS) conducted by the FinnGen consortium and the UK Biobank.

Instrument selection adhered to rigorous criteria: for each protein, we designated the lead cis-pQTL—identified as the single nucleotide polymorphism (SNP) exhibiting the most significant association (p < 5 × 10^□5^) within a 1 Mb region flanking the protein-coding gene—as the primary genetic instrument. To ensure the independence of these instruments, we applied linkage disequilibrium-based clumping with a threshold of R² < 0.001, leveraging the 1000 Genomes Phase 3 European ancestry reference panel and executed via PLINK version 1.9.

#### Protein-Based Prediction Models

##### Cohort and Data Partitioning

The study cohort included all UK Biobank participants with available proteomic measurements, systematically divided into a training set (70%) and a holdout set (30%) for model development and evaluation. Clinical covariates, consistent with those in prior ophthalmic-proteomic analyses, were incorporated as risk factors. This random allocation ensured robust representation across subsets for subsequent predictive modeling.

##### Construction of Proteomic Predictive Models

We constructed predictive models for ocular conditions using XGBoost, a gradient-boosting algorithm, following feature selection with LASSO regression. LASSO, a regularized regression approach, distilled biologically relevant proteins from an initial set of 2,920 by nullifying coefficients of variables with minimal predictive power, effectively managing high-dimensionality and collinearity. The selected proteins were integrated into the XGBoost framework, generating three indices: (1) a proteomic index reflecting protein-specific signals, (2) a clinical index derived from established risk factors, and (3) a combined index merging proteomic and clinical data. Model optimization in the training set employed 10-fold cross-validation, with hyperparameters tuned to maximize the area under the receiver operating characteristic curve (AUC). Predictive performance was assessed in the holdout set, with AUC as the primary metric. Associations between these quintiles and ocular outcomes were quantified in the holdout set using Cox proportional hazards models, providing a comprehensive assessment of predictive accuracy and temporal trends.

##### Unsupervised clustering of ocular phenotypes based on proteomic associations

To identify molecularly coherent groups of ocular phenotypes, we applied hierarchical clustering to a matrix of scaled protein–phenotype associations. Each row represented an ocular phenotype (n = 80; 43 eye diseases and 37 quantitative traits), and each column represented a plasma protein (n = 2,920). The input matrix was constructed using normalized effect estimates (Z-scores) from multivariable-adjusted linear or Cox regression models, depending on the outcome type. Missing values were imputed as zero, reflecting no association signal.

Pairwise distances between ocular phenotypes were computed using Euclidean distance, and clustering was performed using Ward’s minimum variance method (ward.D2) as implemented in the hclust function in R. The optimal number of clusters was determined based on visual inspection of the dendrogram, gap statistic, and biological interpretability. A circular chord diagram was used to visualize the cluster composition and highlight shared membership between diseases and traits. For each cluster, pathway enrichment analysis was performed on cluster-enriched proteins using g:Profiler, and representative biological terms were summarized.

## 4. Statistical Analyses

All statistical tests were two-sided unless otherwise indicated. Analyses were conducted in R (version 4.2.1; R Project for Statistical Computing). Associations between plasma protein concentrations and ophthalmic outcomes, including health ocular traits and incident eye diseases, were evaluated using generalized linear models (GLMs) and Cox proportional hazards regression, as detailed in the respective proteomic-phenotype association subsections. Feature selection for predictive modeling employed LASSO regression via the glmnet package, while XGBoost-based prediction models were implemented using the xgboost package. Model performance was quantified by the area under the receiver operating characteristic curve (AUC) using the pROC package. Enrichment analyses to identify overrepresented biological pathways were performed using the clusterProfiler package. Two-sample Mendelian randomization (MR) analyses were executed using the TwoSampleMR package to infer causal relationships. All packages were compatible with R version 4.2.1.

## Contributors

DL conceptualized and designed the study, completed the statistical analyses, drafted the initial manuscript, and reviewed and revised the manuscript; LC and NW contributed to the conceptualization and design of the study, the statistical analyses and initial drafting of the manuscript, and reviewed and revised the manuscript; HL, XC, XZ, JZ, JW, LC, and NW reviewed the manuscript. The corresponding author attests that all listed authors meet authorship criteria and that no others meeting the criteria have been omitted. All authors approved the final manuscript as submitted and agree to be accountable for all aspects of the work.

## Data Availability

Individual-level phenotypic and proteomic data are available upon application to the UK Biobank (https://www.ukbiobank.ac.uk). GWAS summary statistics were obtained from UK Biobank and the FinnGen consortium, accessible via the FinnGen website (https://www.finngen.fi) and Supplementary files.

## Funding and Acknowledgements

This work was supported by National Natural Science Foundation of China (82130029). This research has been conducted using the UK Biobank Resource under Application Number 515707. We thank the UK Biobank participants and coordinators for their invaluable contributions. Analyses were conducted using the UK Biobank Research Analysis Platform (RAP), developed by DNAnexus in collaboration with the UK Biobank.

